# Chaos, Percolation and the Coronavirus Spread: a two-step model

**DOI:** 10.1101/2020.05.07.20094235

**Authors:** Aldo Bonasera, Hua Zheng

## Abstract

We discuss a two-step model for the rise and decay of a new coronavirus (Severe Acute Respiratory Syndrome-CoV-2) first reported in December 2019, COVID-19. The first stage is well described by the same equation for turbulent flows, population growth and chaotic maps: a small number of infected d_0_ grows exponentially to a saturation value d_∞_. The typical growth λ time (aggressive spreading of the virus) is given by 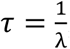, where *λ* is the Lyapunov exponent.!After a time t_crit_ determined by social distancing and/or other measures, the spread decreases exponentially as for nuclear decays and non-chaotic maps. Some countries, like China, S. Korea and Italy are in this second stage while others including the USA are near the end of the growth stage. The model predicts 15,000 (±2,250) casualties for the Lombardy region (Italy) at the end of the spreading around May 10,2020. Without the quarantine, the casualties would have been more than 50,000, hundred days after the start of the pandemic. The data from the 50 US states are of very poor quality because of an extremely late and confused response to the pandemic, resulting unfortunately in a large number of casualties, more than 70,000 on May 6, 2020. S. Korea, notwithstanding the high population density (511/km^2^) and the closeness to China, responded best to the pandemic with 255 deceased as of May 6,2020.

## Introduction

Chaotic models have been successfully applied to a large variety of phenomena in physics, economics, medicine and other fields [1-6]. In recent papers [7,8] a model based on turbulent flows and chaotic maps has been applied to the spread of COVID-19 [9]. The model has successfully predicted the rise and saturation of the spreading in terms of probabilities, i.e. the number of infected (or deceased) persons divided by the total number of tests performed. Also a dependence on the number of cases on the population density has been suggested [7] and the different number of fatalities recorded in different countries (or regions of the same country) was attributed to hospitals overcrowding [8]. In this paper we would like to extend the model to the second stage, i.e. the decrease of the number of events due to quarantine or other measures [10]. Different fitting parameters of the model are due to the different actions, social behaviors [11], population densities [7], pollution [12] etc. of each country but there are some features in common and it is opportune to first have a look to some data available on May 6,2020 and updated in this revised version of the paper to the end of June, 2020.

In the figure 1, we plot the number of positive (top panels) and deceased (bottom panels) as function of time in days from the beginning of the recordings. Some data have been shifted along the abscissa to demonstrate the similar behavior. Different countries are indicated in the figure insets. As we can see all the EU countries display a very similar behavior including the U.K. notwithstanding the Brexit. The USA case has been shifted of 38 days, which is the delay in the response to the pandemic resulting in the large number of fatalities. In contrast, S. Korea reacted promptly and was able to keep the number of positives and more importantly the death rate down. Among the EU countries, Germany shows the lowest number of deceased cases, which could be due to different ways of counting (for instance performing autopsies to check for the virus like in Italy). In any case, the analysis in ref.[8] shows that different regions of Italy have lower mortality rates (for instance the Veneto region which borders the Lombardy region-the highest hit) compatible to Germany. Thus, similar to [8], we can assume that different overcrowding of health facilities, retirement homes, jails etc. might be the cause [11,12] for the differences displayed in the figure 1. The striking feature in figure 1 is that all countries seem to have reached saturation while the USA is still growing both as the number of positive and deceased. Notice the striking similarity of figure 1 with the model of population growth as originally proposed by P.-F. Verhulst [13].

**Figure 1.**
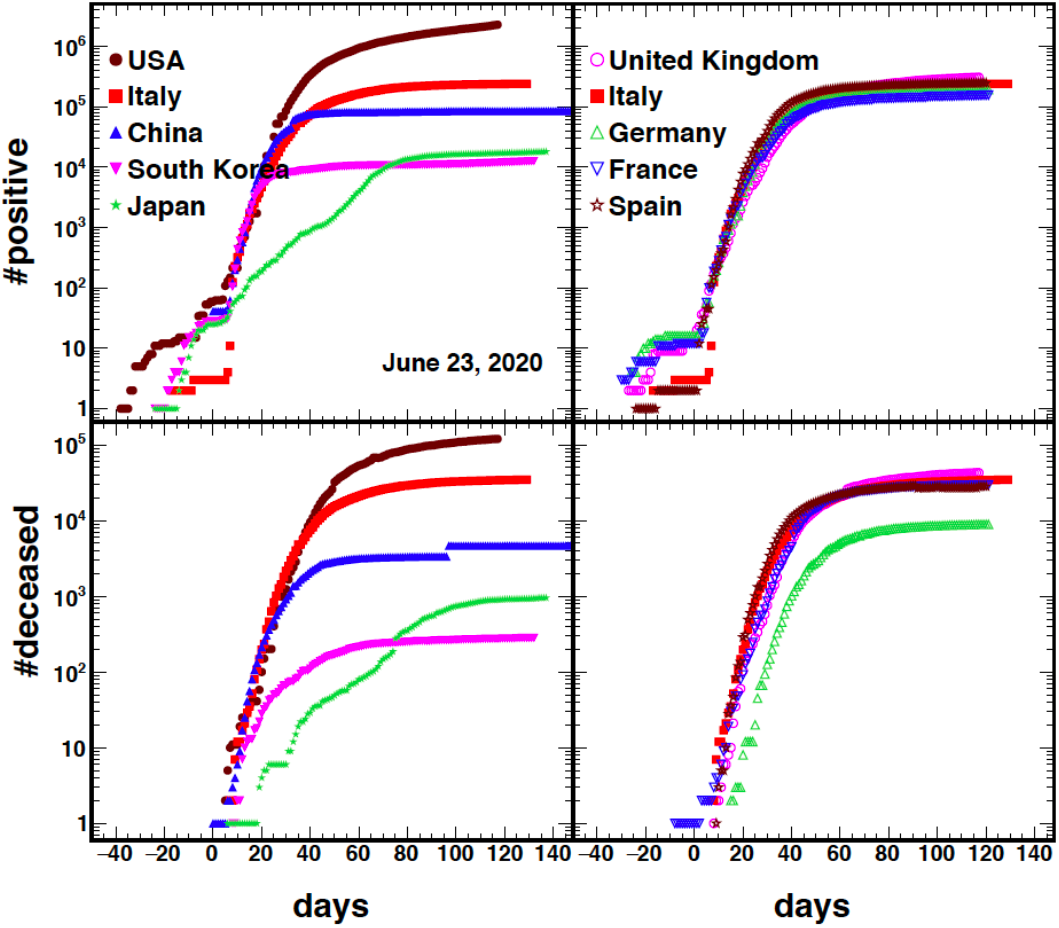
Number of positive (top panels) and deceased (bottom panels) as function of time for different countries indicated in the inset. Time t=0 was suitably chosen to match the exponential growth for the number of positive and it was kept the same for all the other plots, figures 1,3.

To contrast the pandemic, many countries have adopted very strict quarantine measures. Social distancing and other measures decrease [8,11,12] the probability to remain infected thus we expect that countries with lower population density might have better and faster success. On the other hand, if some countries adopt non-effective measures or it is too late in the response, the lower population density might hinder the problem for some time. Thus in order to better stress the efficacy of the quarantine, we have plotted in figure 2 the number of cases DIVIDED by the population density, assuming that it is much easier to perform social distancing if the population density is low. In the figure 2 we see that S. Korea and Japan, even though their densities are rather high, 511/km^2^ and 334/km^2^ respectively, perform best. We should also consider that S. Korea (or Japan) is ‘*across the street*’ from China, the epicenter of the infection [7,8], while the other countries are located across a continent or an ocean giving further advantages to organize a response which unfortunately turned out to be weak and badly organized. The last data points for China reflects an adjustment to the death rate in Wuhan, which probably had similar problems like the Lombardy region in Italy [8]: we will not be surprised to see future corrections.

**Figure 2.**
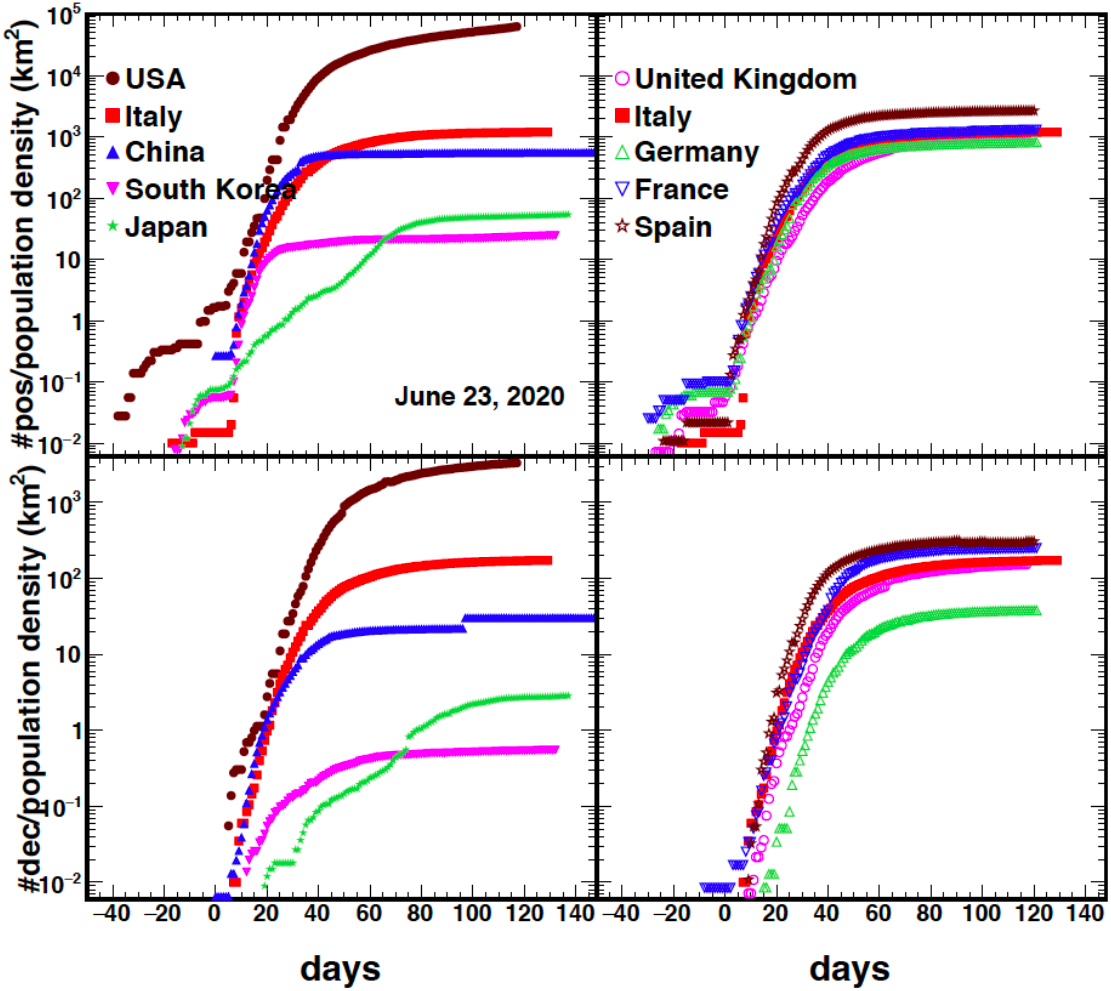
Number of cases divided by the population density of each country vs time, compare to figure 1.

There is a ‘hidden’ parameter in the figures 1 and 2: the number of tests performed daily. Zero tests, zero cases and no problem but then the hospitals get filled with sick people and we have a pandemic. In order to have realistic information on the time development of the virus, it is better to calculate the total number of cases DIVIDED by the total number of tests, this defines the probability to be infected or the death rate probability due to the virus. We stress that such probability may be biased since often the number of tests is small and administrated to people which are hospitalized or show strong signs of the virus [7,8]. The values we will derive must be considered as upper limits but the time evolution should be realistic.

Not all the countries provide the number of tests performed daily (China). In the figure 3, we plot the probabilities vs time for the same countries as in the figures 1 and 2. As we can see some cases show a smooth behavior indicating prompt and meaningful data taking. Large fluctuations or missing data are also seen at the beginning, which means that the number of early daily tests was very small. All countries show a decreasing behavior at long times both for the positive and deceased cases suggesting that the pandemic is getting under control, but with different rates. S. Korea and Japan display a similar behavior but with much lower values. Germany ‘performs best’ among the EU countries analyzed here most importantly regarding the death rate. The USA, which was showing an increasing trend in the figures 1,2, displays a decreasing probability but at a lower rate and the increase in the previous figures may be attributed to the increase in the test number.

**Figure 3.**
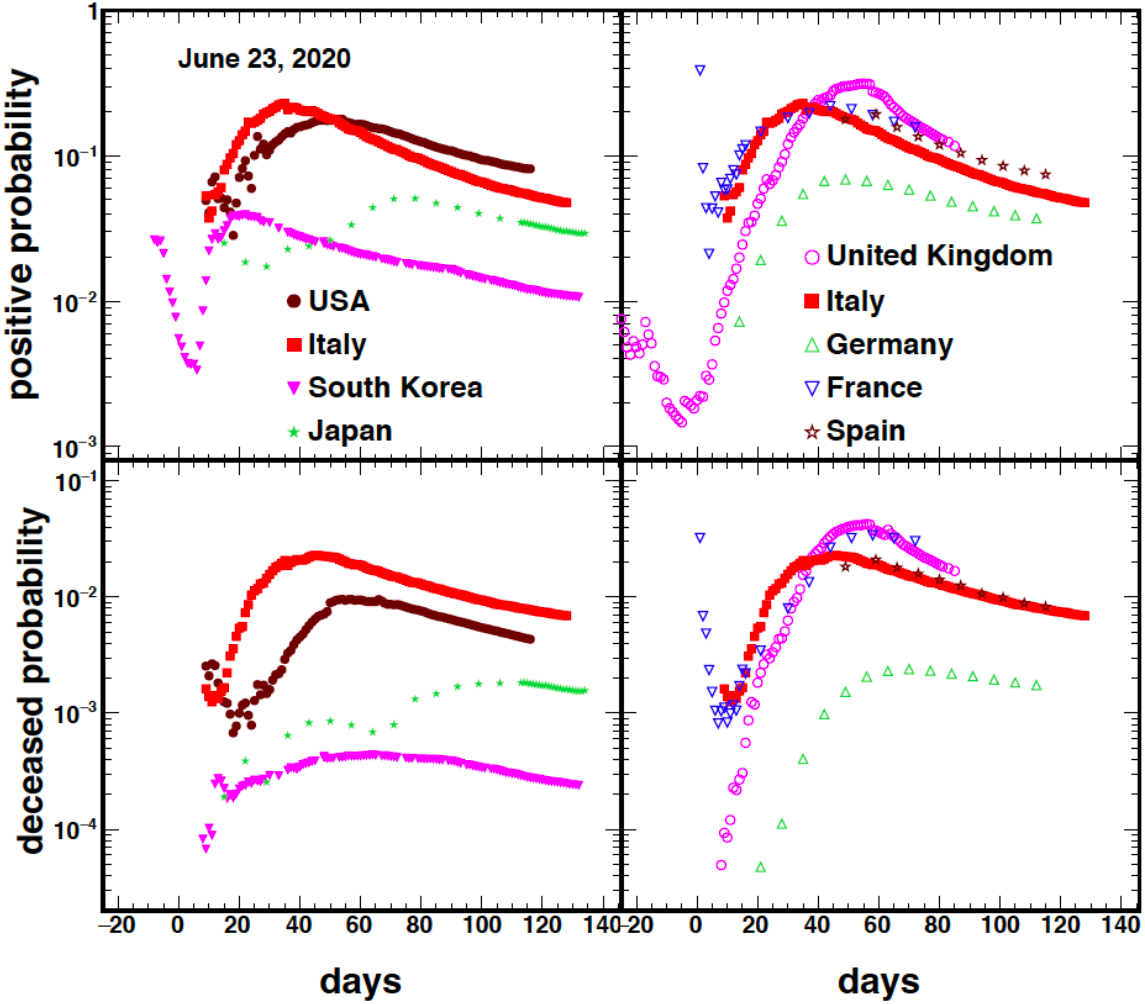
Probabilities vs time for the countries indicated in the inset. Some countries stopped providing the number of tests performed daily (France on May 5^th^ and the UK on May 22^nd^), other countries are providing this information periodically (Spain, Germany).

### The Model

We have discussed and applied the first stage of the model in refs.[7,8]. We briefly recall it and write the number of people (or the probability) positives to the virus (or deceased for the same reason) as [1-8,13]:

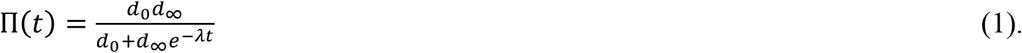

In the equation, t gives the time, in days, from the starting of the pandemic, or the time from the beginning of the tests for the virus. At time t=0, ゠(0)=d_0_ which is the very small value (or group of people) from which the infection started. In the opposite limit, *t* → ∞, Π ∞ = *d*_∞_, the final number of affected people by the virus. Equation (1) has the same form observed in the figures 1 and 2, but in reality it should be applied not the number of positives (or deceased) but to their probabilities, i.e. the number of cases divided by the total number of tests. The main reason for this definition is to avoid the spurious time dependence due to the total number of tests, which varies on a daily basis and very often not in a smooth way [7,8]. In the figure 3 we have plotted the probabilities for different countries since the data are available. It is important to stress that the information on the total number of daily tests is crucial and should be provided also to avoid suspects on data handling. If we treat equation (1) as a probability then we expect to saturate to *d*_∞_ at time *t*_crit_. At later times, if social distancing is having an effect, we expect the probability to decrease and eventually tend to zero. In the figure 4, we see exactly such a behavior for the cases of two Italian regions: Lombardy and Sardinia [8], https://github.com/pcm-dpc/COVID-19. For times larger than t_crit_ the decrease is exponential and can be described as for nuclear decays and non-chaotic maps [1,10]:

**Figure 4.**
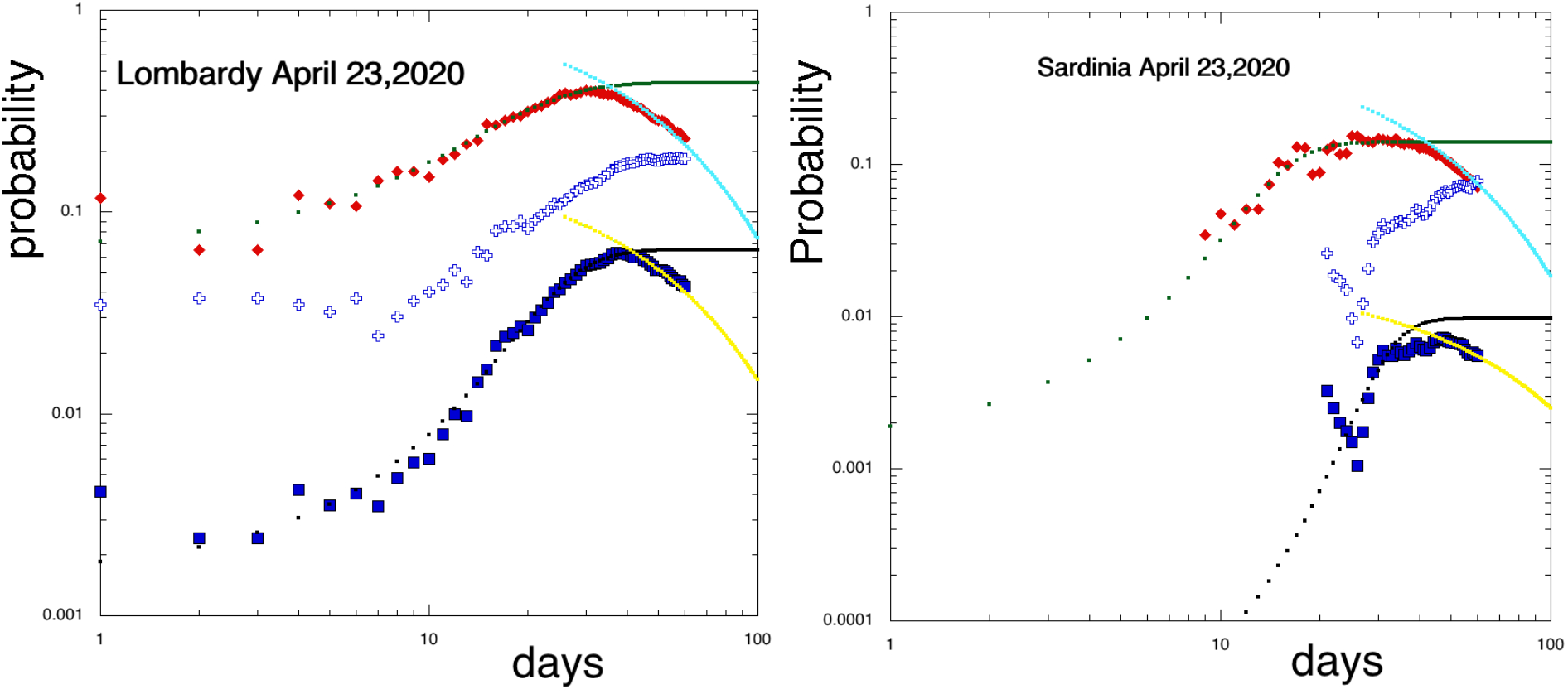
Probability for positive (rhomb symbols) or deceased (square symbols) vs time in days for the Lombardy (left) and Sardinia (right) regions. The open crosses give the ratio positive/deceased and reach almost 20% for Lombardy [8]. The continuous points are obtained from eq.(1) and the exponential decay from eq.(2). Updated data are given in figure A1.

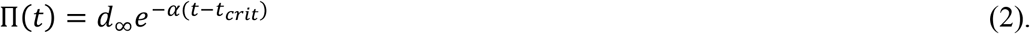

*α* and t_crit_ are fitting parameters. Values for the Lombardy region are *α* = 0.0268(0.025) d^-1^ and t_crit_=39(42)d for the positives (deceased). We can infer the decay time as *τ*_d_=1/α=37(40)d suggesting that roughly *τ*_d_ after the maximum the pandemic should be over, i.e t_max_≈ t_crit_+ *τ*_d_=76(82) days from February 24,2020. To demonstrate the predictive power of the model we updated the data to the Lombardy case in the appendix, figure A1. A similar agreement to other Italian regions is found [8].

From the figures 4 and A1, it is quite easy to derive the value of t_crit_ given by the maximum. This value differs slightly for the positive and the deceased as well as for the different regions. Thus it is important to have enough data to perform best fits using equations (1) and (2). The value of t_crit_ depends on many factors including the population density, the weather temperature, humidity etc. and especially social distancing or any other measures used to contrast the pandemic. If no measures are adopted (herd immunization or natural selection approach), such as for some countries like Sweden (and the UK at first), then we expect the plateaus in figures 3 and 4 to last longer but eventually the process will be described by equations (1) and (2). The herd immunization approach might be reasonable if we do not think we are going to get a vaccine soon. However, in such cases we may also expect to be flooded by positives and deceased persons jeopardizing the health structures and harm the sanitary personnel [8]. A country like Sweden with excellent sanitary structures and low population density (25/km^2^) may succeed in this task, but the same attempt in the UK (279/km^2^) was a disaster and quickly abandoned as can be seen from the figures 1-3. In particular in figure 3 we see that the UK have the largest probabilities, https://www.who.int/emergencies/diseases/novel-coronavirus-2019. Of course the predictions have validity if the conditions are not changed, for instance relaxing the quarantine too soon. If these conditions are modified then we may have a rapid increase of the cases again and return to the original curve given by equation (1), such a behavior might be noted in figure 3 for Japan. At the same time when the Olympics 2020 were under discussion, Japan interrupted the COVID-19 testing as can be seen from the plateaus in figures 1 and 2 lasting approximately 15 days. Thus it is important to understand when to relax the measures and for this reason we have plotted in figure 4 two cases. Lombardy is the worst case in Italy with more than 14,000 deceased in contrast to Sardinia with about 100 as of May 6,2020. In the figures we can see that the probabilities are much lower for Sardinia, which could be regarded in some sense as the future of what should eventually happen in Lombardy because of the quarantine. The population density of Sardinia is relatively low, 69/km^2^, and it is an island away from the mainland. This situation is in many respects very similar to S. Korea with lower population density. Thus, the measures might be relaxed in Sardinia following the example of S. Korea after careful instructions to the population and random every day testing to search for positive and isolate them. This will provide crucial information on the social behavior and on the virus spread. We will show below that the model predicts a small number of positive and deceased for Lombardy around or after May 10,2020 thus shelter at home might be extended up to that day. It would be important to send some signals to the population of return to normality after months of sheltering by organizing for example sportive events in Sardinia. The Italian national sport, “Serie A”, might organize 2-3 games per day in different Sardinian towns, with empty stadiums and broadcasted live. Other limited activities but strongly controlled could be allowed in less affected regions such as Calabria, Abruzzo and other southern Italian regions discussed in ref.[8]. Releasing all measures for the entire country at the same time might be not too wise. Looking at other countries experiences, we would suggest that quarantine should not be released before the probability for positives is less than 4% (the maximum of S. Korea, figure 3). Below such a value, the other countries may follow the S. Korean approach but if they are not organized to do that, reopening too soon may be dangerous.

The model describes very well the data and might be used for the everyday control on the resurgence of the pandemic. It offers another great advantage: we have described a way to eliminate misleading inputs due to the number of everyday test. We can proceed in the inverse direction in order to predict the total number of deceased and positives cases. The task that we have now is much easier and it is the prediction of the daily tests for each case. As we have seen from figures 1-3, there were some wrong decisions taken by the different countries at the beginning of the pandemic (apart S. Korea and Japan) resulting in a very small number of tests. After 1-2 weeks the number of test per day was increased and eventually become constant. It is this behavior we have to predict in order to extend our model to the total number of cases. In figure 5 we plot the total number of tests vs time from the beginning of the recordings for Lombardy (February 24,2020). We have fitted the data with a power law function as indicated in the figure but any other suitable function might do as well. As we see from the figure, the Italian data is fitted very well with a small error on the fitting. Fits performed to other countries give a power exponent ranging from 0.73 (S. Korea) to 4.1 (UK). This is also an indication of how well organized the response to the pandemic is. In the ideal case we expect the power to be about 1, the value for the UK suggests some change of strategy (from i.e. herd immunization to quarantine) and because of such high value we are not able to make predictions on the total number of tests say 50 days after May 6,2020. In this revised version, the new data for the UK-see figure 3, allows us to make predictions as discussed below.

**Figure 5.**
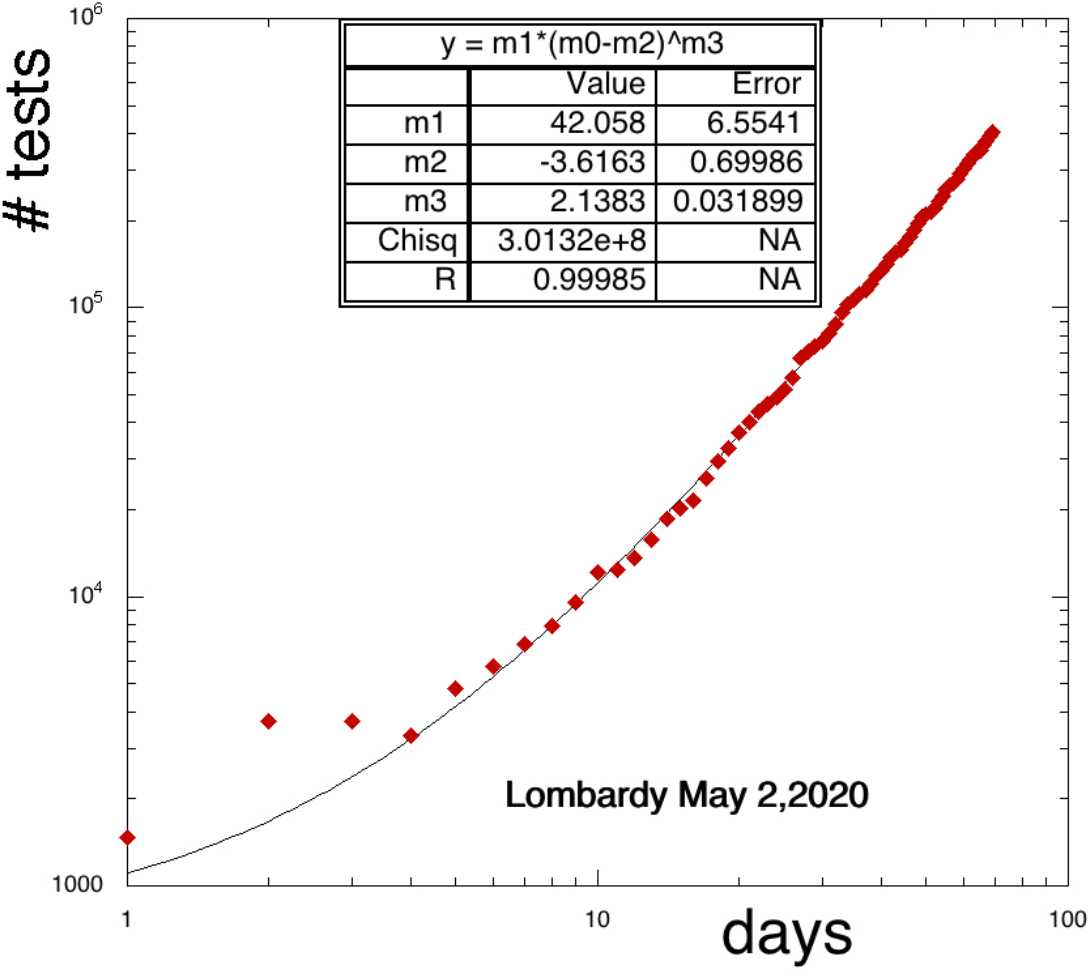
Total number of tests as function of time for Lombardy. The fitting function and its values are displayed in the insets.

Multiplying equations (1) or (2) by the predicted number of tests from figure 5, gives the total number of predicted cases and are compared to the data in figure 6. We assume a conservative 15% error in our estimates due to the different fit functions. Mostly important, an error is coming from laboratory testing with current methods [14-19]. Without social distancing, using equation (1) gives 360,000 (±54,000) for the positives and 53,000 (±7,950) for the deceased 100 days after the beginning of the pandemic in Lombardy. If the exponential decay given by equation (2) is taken into account (due to the quarantine), the values decrease to 80,000 (±12,000) and 15,000 (±2,250) respectively, thus about 38,000 saved lives in Lombardy alone! There is an important difference between the two stages: if the first stage alone would be at play, the pandemic may continue after the 100 days and eventually slow down at longer times. Recall that the Spanish flu started in 1918 and lasted almost 36 months with an enormous death toll, https://www.washingtonpost.com/graphics/2020/local/retropolis/coronavirus-deadliest-pandemics/.

**Figure 6.**
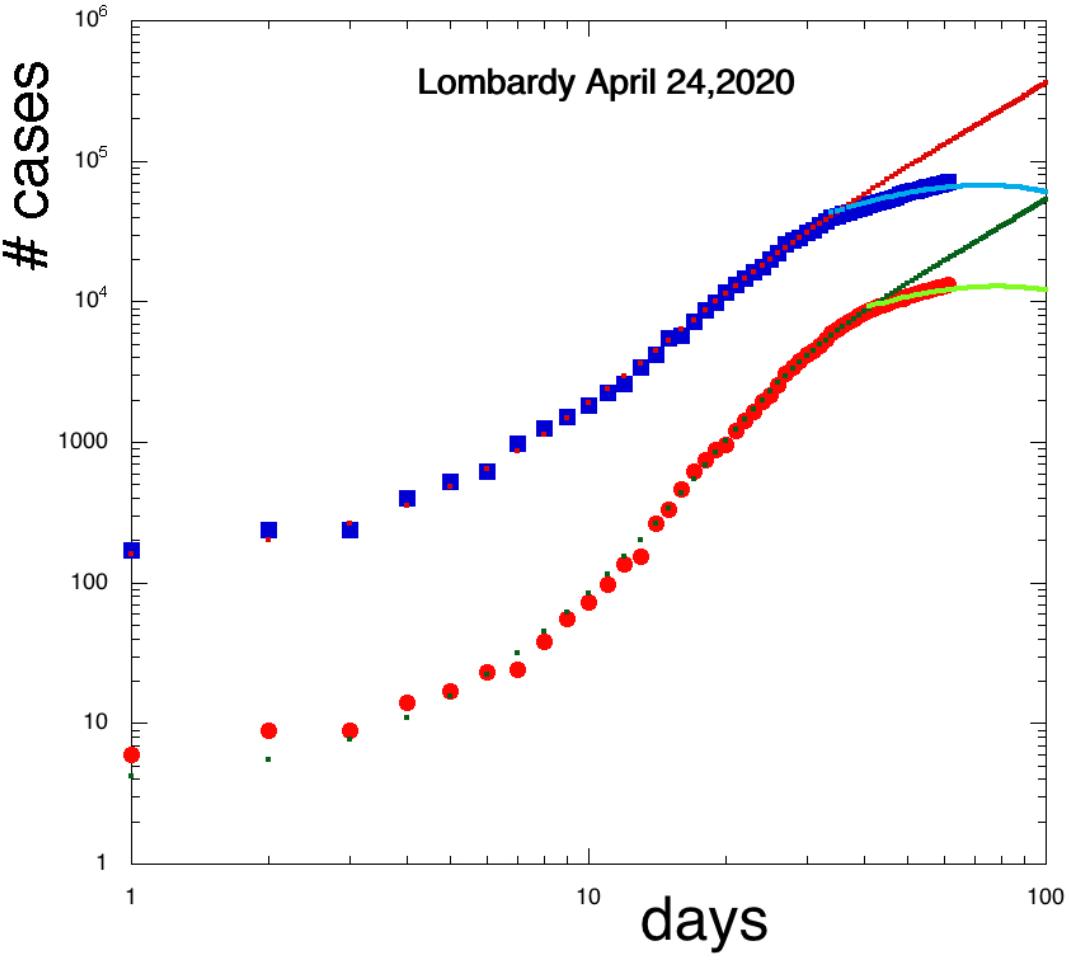
Predicted cases with and without quarantine as function of time, see text. Data for positive and deceased are given by the square and circle symbols respectively.

Because of the second stage, now the predicted values are given by the maxima in the figure 6, these occur 76 and 82 days respectively after the start of the pandemic recording, i.e. May 10 and 16,2020 respectively. These values are close to the sum of t_crit_ and *τ*_d_ reported above. If we assume a power law to reproduce the available data for the number of tests, figure 5, then we can write the total number of cases in the second stage as:

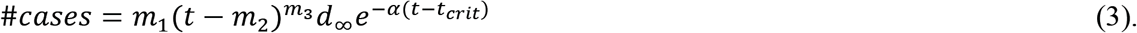

The fitting parameters m_1-3_ are reported in figure 5 for Lombardy. To find the maximum ofM equation 3, we simply equate its derivative to zero:

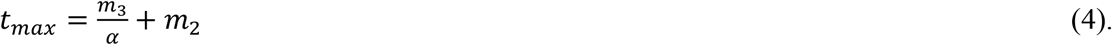

Using the empirical relation above connecting t_max_ and t_crit_ we get:

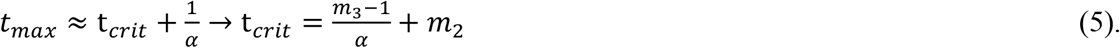

This relation is very useful especially when the data does not show the exponential decrease since it reduces the number of free parameters entering equation (2). Similar relations can be derived for different parameterizations for the total number of tests.

### The US states

Figures 1-3 show that the USA was hit hard by the COVID-19 resulting in different responses from the different states. In this section we will analyze some of these states and more analyses can be found in the supplemental material or available from the authors. In figure 7, we plot the probabilities for the state of California (Ca) for the period indicated in the inset, compare to figure 3 and 4. The discontinuities are due to the change in the number of tests performed daily. Notice that March 14,2020 coincides with the quarantine declaration in Italy, thus it was not a surprise that the virus spread quickly. Fortunately, the San Francisco mayor and the California governor placed strict restriction as early as March 6 without waiting for better testing, https://www.sfdph.org/dph/alerts/files/HealthOfficerLocalEmergencyDeclaration-03062020.pdf. This action saved a large number of lives and kept the ratio deceased/positives very low, compare to figure 4. We can correct in some cases for the low number of tests. Large data taking has a better statistical value thus we renormalize the data where the jumps occur to the value at later times. In the right panel we display the result of the renormalization together with the fit using equation (1).

**Figure 7.**
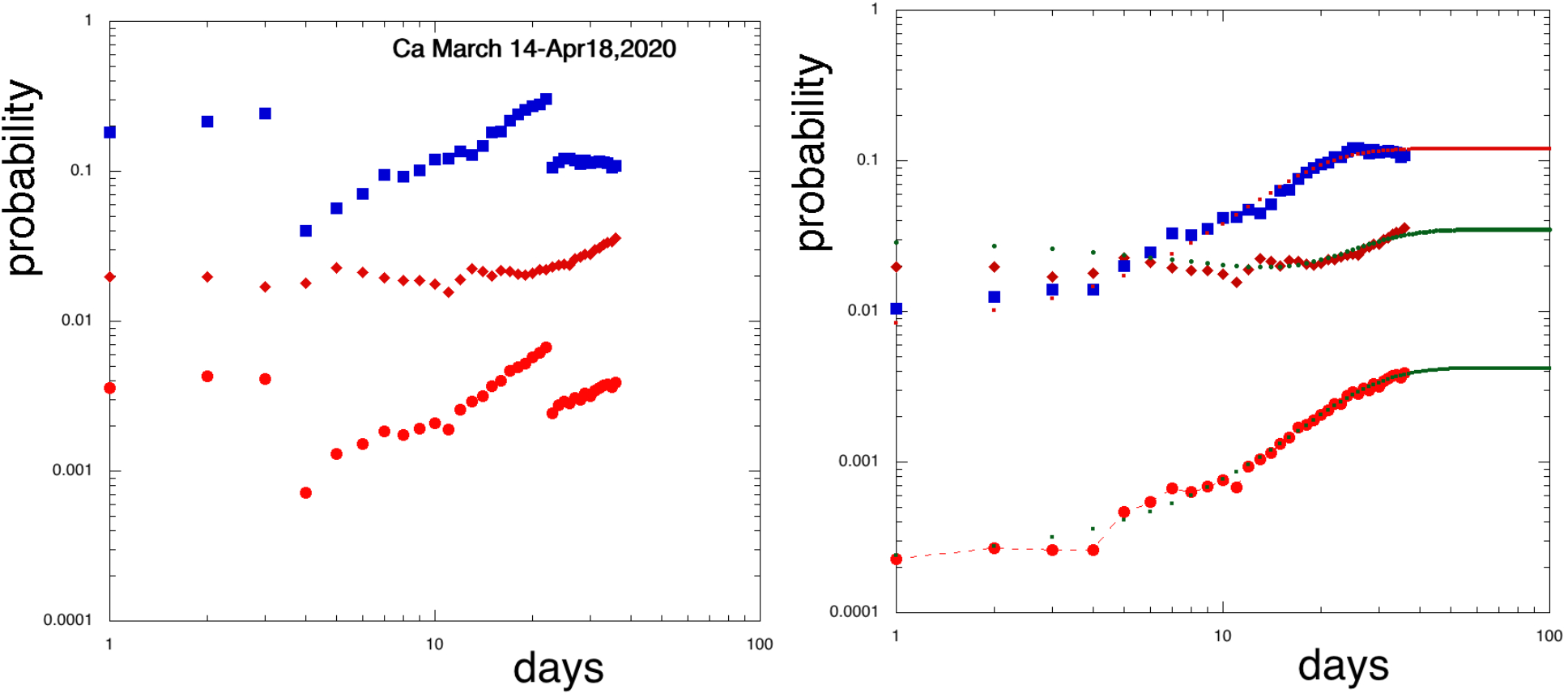
Positive (square symbols) and deceased (circle symbols) probabilities vs time in days. The rhomb symbols represent the ratio deceased/positive independent on the number of tests. The right panel is obtained after renormalization, see text.

The hardest hit state was New York. In the figure 8 we display the probabilities together with the fits using equations (1) and (2), compare to figures 4,7. The ratio deceased/positives seems smaller than the Lombardy one, however particular attention should be paid to the counting methods and some confusion might arise if the data refer to the state of New York (NY), https://coronavirus.jhu.edu/map.html, or to New York city (NYC), https://covidtracking.com/data/state/new-york#historical, the difference being roughly 5000 deaths since most cases are in NYC. The bending down of the curve is evident and we can make a prediction using equation (2). The resulting fit is displayed in figure 8, it follows well the available points but further confirmation will be given by future data. In the appendix, figure A2, we compare the data available on June 23,2020 to the model confirming its validity.

**Figure 8.**
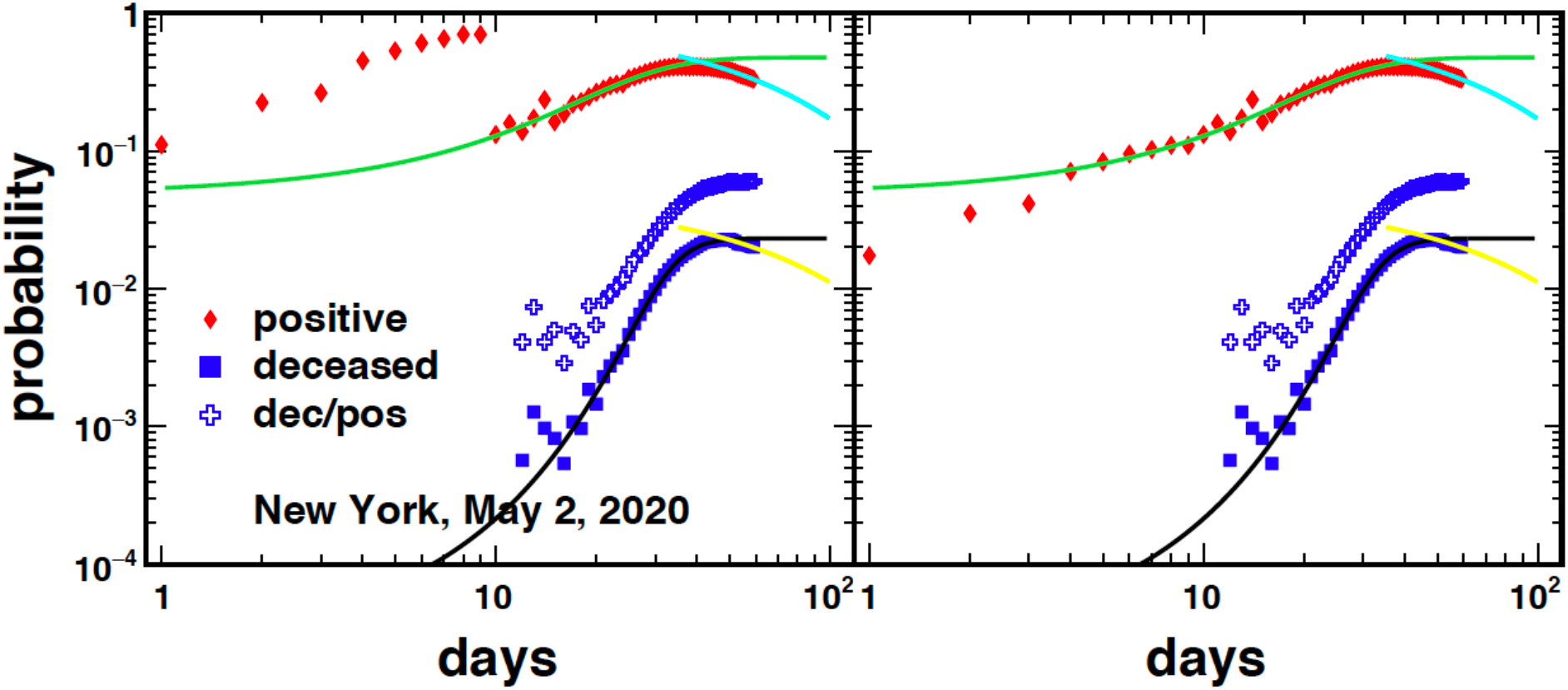
Same as figure 7 for the state of New York. See figure A2 for an update.

Using the predicted number of tests for NY given in figure 9, left panel, and the probability fits from equation (1) and (2) displayed in figure 8, we can predict the total number of cases as for Lombardy. The results are plotted in figure 9, right panel, for the first 100 days from the start of the recordings.

**Figure 9.**
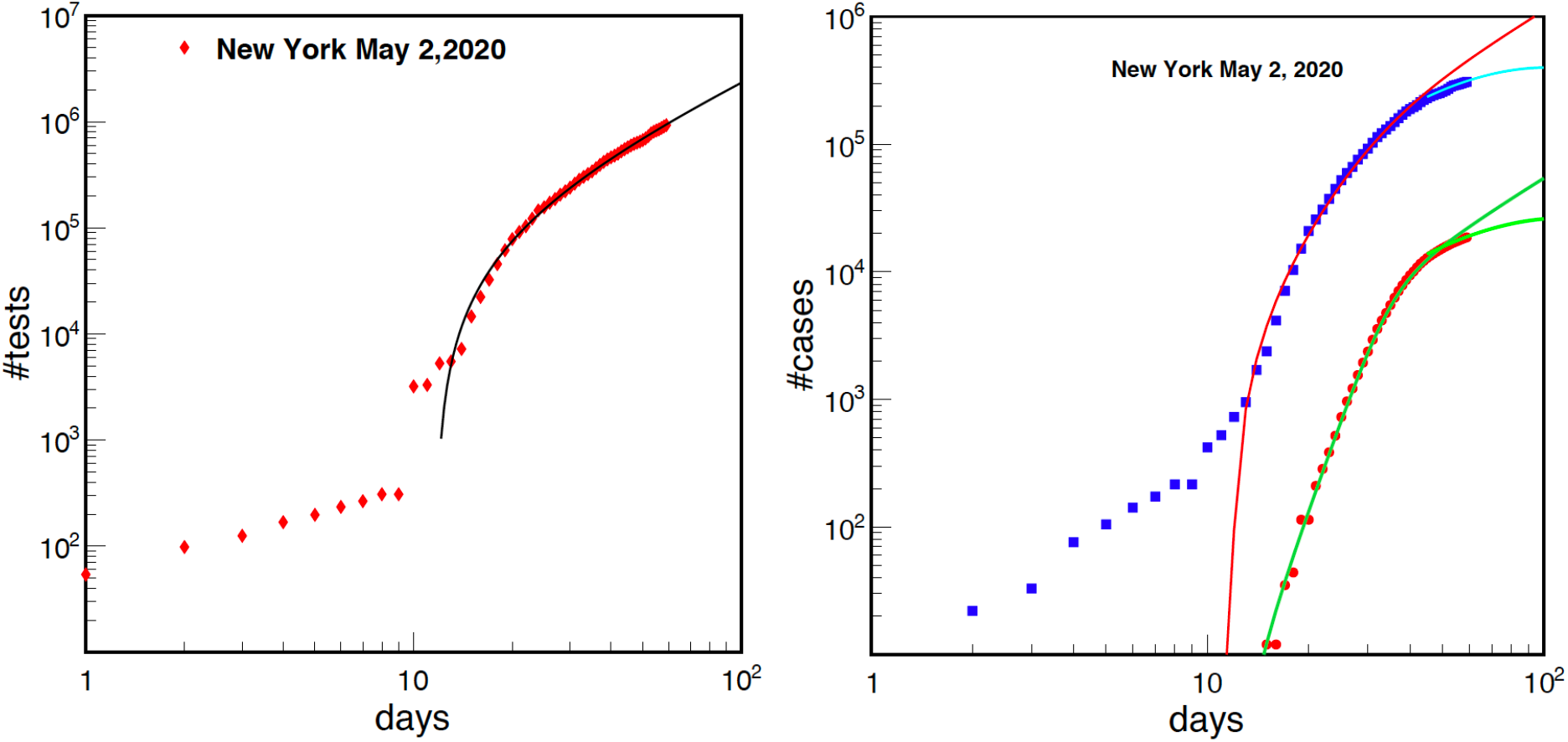
Same as figures 5 and 6 for the state of New York.

## Conclusions and outlook

We have proposed a two-step model to the rise and decay of the pandemic due to the COVID-19. The model needs some input parameters to predict the time evolution up to the saturation of the probability as in equation (1). Once the plateau is reached, given by the d_∞_ parameter, the probability remains constant for some time depending on the quarantine measures or other environmental factors [11,12]. For the Italian case, the first test was published on February 24 and equation (1) was fitted on March 10 before the quarantine was announced, i.e. March 14 [7]. The plateau was reached around March 24 as predicted by the model. These dates suggest that the quarantine was not effective in reducing the maximum probability and the time when this was reached. The quarantine became effective roughly 10 days after saturation. Thus we can estimate that it takes more than 3 weeks before the quarantine gives an effect and the probabilities start decreasing, this is the value of t_crit_ entering equation (2). We can assume that if the quarantine was announced say 14 days earlier, then the exponential decrease, equation (2) would had intercepted the rise, equation (1), earlier resulting in smaller probabilities. This is what happened to S. Korea and Japan, and it would explain the differences among countries: the later and the more feeble the quarantine the higher the probabilities and the longer the time it takes to return to (quasi) normality. From these considerations we can estimate the time it takes for other countries also if the probability decrease is not seen yet. After reaching the top of the probability, see figure 3, it took roughly 10 days for Italy to see the decrease. In figure 10 we plot the predicted total number of positive (top panel) and the deceased (bottom panel). Countries, which did not provide the number of daily tests (Spain, China), were not analyzed including the UK because of the large increase in testing especially at later times in coincidence to their Prime Minister hospitalization. In the revised version some cases were added (UK) and updated to June 23,2020.

**Figure 10.**
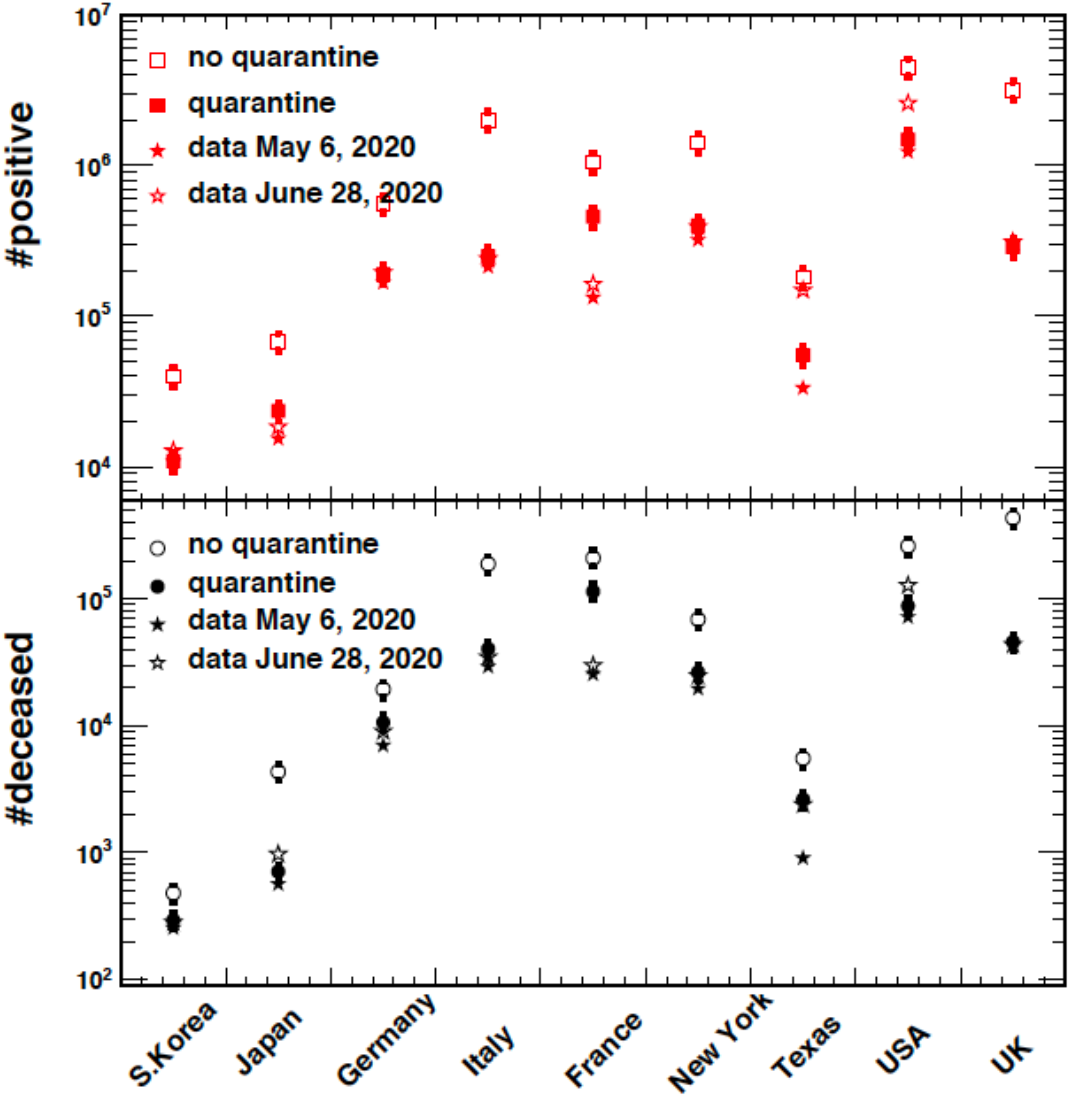
Number of positive (top panel) and deceased (bottom panel) for the countries indicated on the abscissa. Predictions without quarantine measures refer to June 28,2020. The numerical data are reported in table AI.

From the updated data we may notice the overall good agreement to the model with some exceptions so far. France is the most notable with the model over-predicting the data. The reason is the fact that France does not provide data on testing since May 5,2020 when the data was near the highest probability, figure 3. The model under-predicts the USA case while is in good agreement with the states of Texas (for the deceased) and New York. The problem is that the USA did not react to the pandemic as a whole but state by state, this explains why the prediction for the deceased in Texas is good while the positive is underestimated: reopening too early! Recall that there is a time delay between the positive and the deceased. Very soon economic and political reasons influenced heavily the response. As an example, wearing a mask when in public became a political problem [11], which would be ridiculous if not tragic [20]. In the figures A3 and A4 we discuss different cases for different party affiliations for each US state.

Sweden decided to follow a different path of non-imposing the quarantine (herd immunization or natural selection), a choice that could be justified under the assumption that the vaccine will not be available soon enough. In figure 11 we plot the probabilities for Sweden, Finland and Norway since they are bordering countries. The probabilities are quite different especially regarding the death rate. We predict for Sweden about 74,750 (±1.1e4), 8,195 (±1230) positives and deceased respectively on June 28,2020. Since there is no quarantine we are not able to estimate t_crit_ and the decay rate. On the same day, using equation (1), we predict for the other countries the values 18,271 (±2.7e3) and 1,495 (±224) for Finland, 15,225 (±2.3e3) and 391 (±59) for Norway. Thus we see that herd immunization takes a heavy toll not justified by the larger Sweden population (a factor of 2 respect to the other countries considered) and it will be very difficult to explain this choice to the relatives of the victims and their lawyers. We do not have any explanation for the difference in the number of deceased for Norway and Finland since the number of positives is practically the same. Authorities of those countries should investigate this difference further. In figure A5 we update the results to June 23,2020, see also table AII.

**Figure 11.**
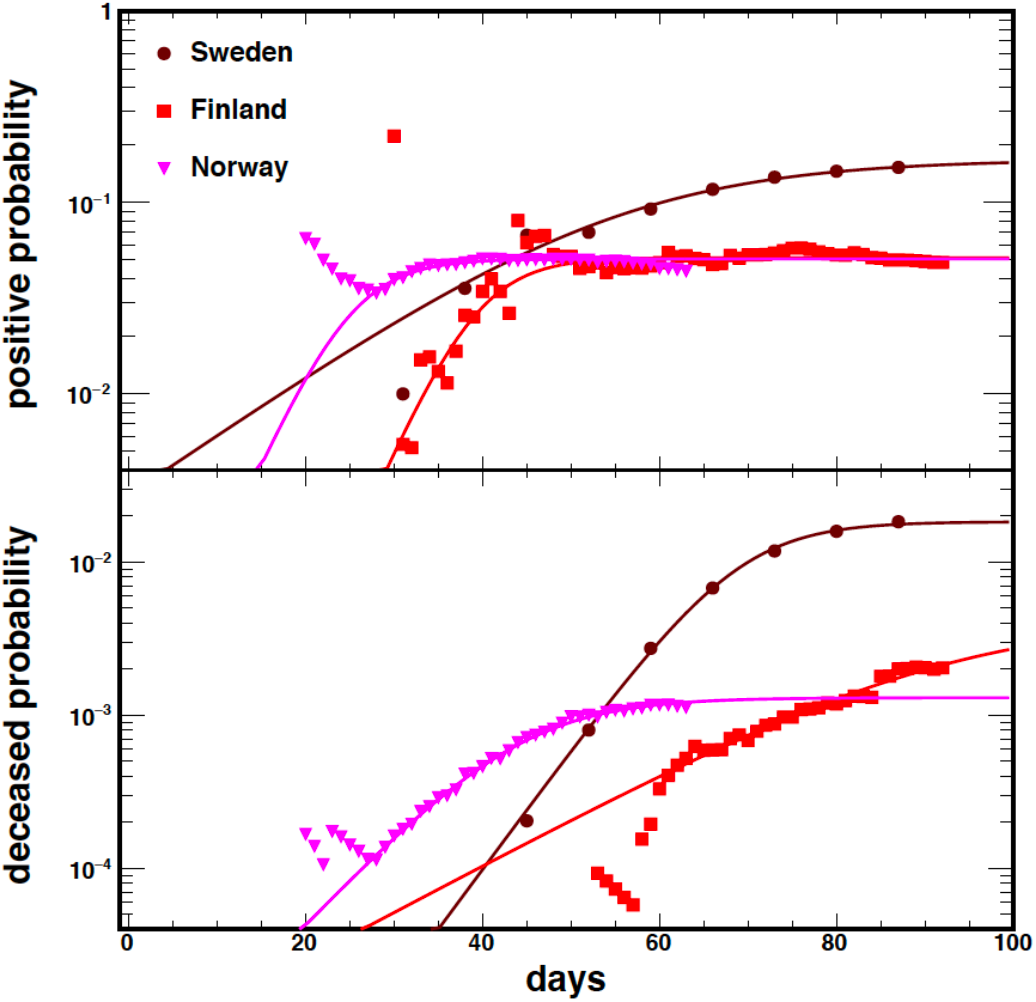
Probabilities as function of time for the countries indicated in the inset. Sweden is adopting the natural selection option resulting in higher probabilities compared to nearby countries. Different starting data depend on which day the complete information needed for the plot was released. Updates are given in figure A5.

One feature worth noticing from figure 11 is the time delay and the slow spread of the Covid-19, this could be due to the extremely cold weather in the winter and early spring for these countries. There is some hope and common believe that the warmer season will help to normalize the situation, as for flu. Other reasons might be put forward, for instance if the virus is somehow adapted to bats, we can naively assume that it will be more deadly for temperatures higher than 10 ^°^C, since below such value most bats hibernate. Temperature difference might explain the spread delay in countries like France, UK and Germany respect to Italy. Of course, other ingredients must be considered such as people flows from/to infected places, population density etc.[11,12]. No matter what the reasons may be for a temperature dependence of the spread it is clear that some systems perform better if it is not too hot or too cold. We can test these hypotheses using the 50 US states data since they cover a wide range of temperatures in the spring season. In the figure 12, we display the results obtained using the data on May 3,2020. Different states values were averaged if their temperatures differ about 1^°^C in order to have better statistics. Even though a smooth behavior is not observed we enforced (optimistic) Gaussian fits, which give 10^°^ C at the maximum and a similar variance, see the inset in the figure 12. The large error bars and the discrepancies respect to the fit at higher temperatures refer to touristic places: Florida, Hawaii and Louisiana, particularly popular during spring break. If we take this result at face value, it predicts about 240 people per million inhabitants to be positive to the virus in the summer with 35 ^°^C average temperatures. Even if this value seems small, it is a seed d_0_ to restart the pandemic. We can already see this in the figure 12 from the large increases over the Gaussian fit corresponding to the high population density states and large touristic flows. It might suggest that low temperatures in hospitals may decrease the virus aggressive spreading, keeping in mind that a vaccine is the only definitive solution. Until then we can only aggressively test and isolate positives similarly to the S. Korean approach to the pandemic.

**Figure 12.**
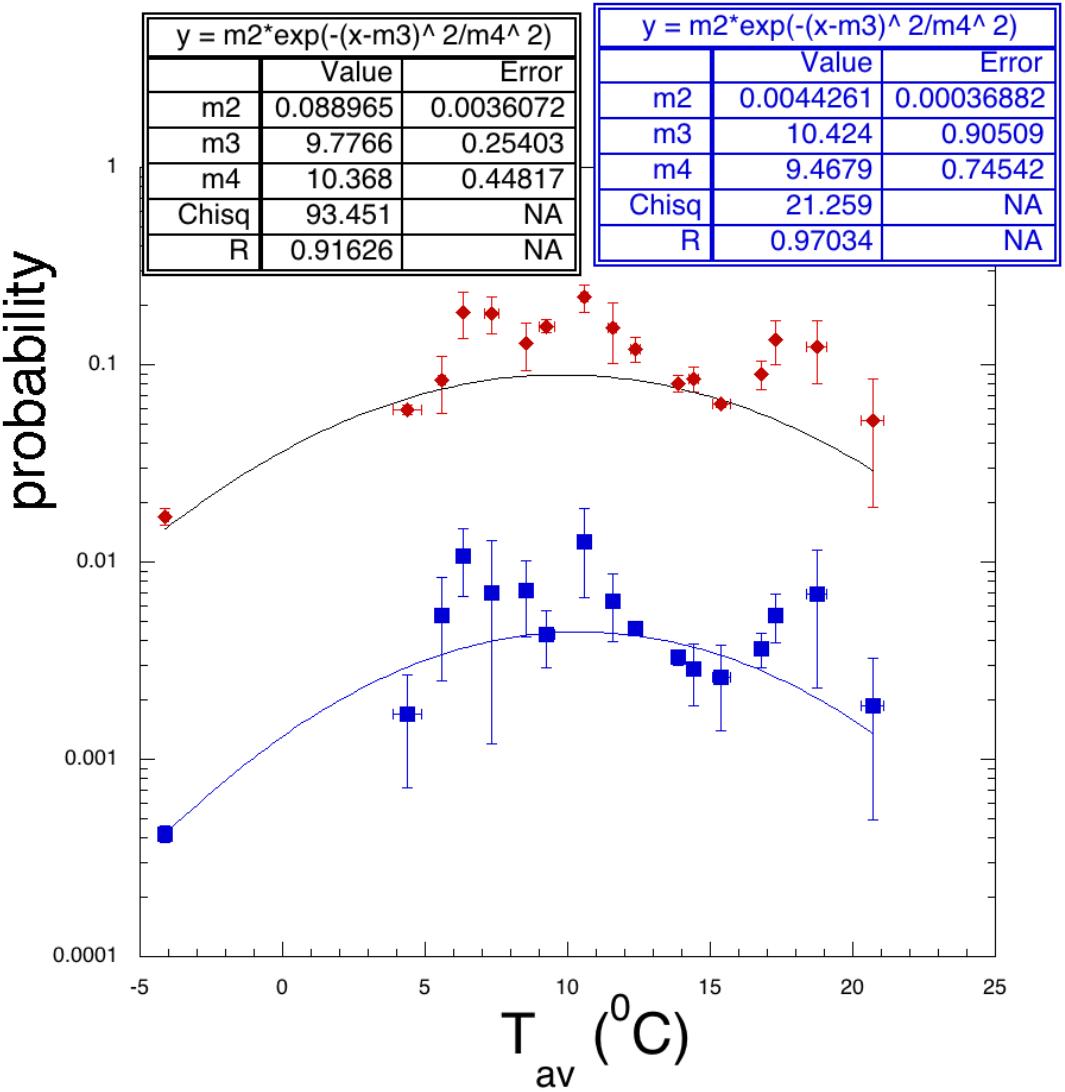
Positive (rhomb symbols) and deceased (square symbols) probabilities vs average temperature of the US states in the spring season. The data to calculate the probabilities were collected on May 3,2020. To get better statistics, averages were performed over states differing about 1 ^°^C. Enforced Gaussian fits are also included and the fit parameters given in the inset.

The preceding discussion and figure 12 suggests that the ‘common believe’ that the summer season weakens the virus is (unfortunately) not supported by the data. Another popular argument widely discussed in the press [21] is the beginning time of the virus spread. The first reported cases to the World Health Organization date December 31, 2019 (hence the name Covid-19 [11]) from the Wuhan region in China. In ref.[7], the Wuhan case was analyzed and concluded that the time it took for the virus to peak is of the order of 15 days, thus around the middle of January 2020. We have seen the devastating effect of the virus on the society with tens of thousand positive and deceased in short times, thus to hide the pandemic is impossible. China does not provide the number of daily tests and we cannot derive the typical times entering our model as discussed in section 2. However, we can derive these times for most of the countries/states/regions discussed in the paper. In figure 13, we plot t_max_ & t_crit_ vs 1/λ (the Lyapunov time) and for completeness the corresponding numerical values are reported in table AIII. The Lyapunov time gives the rate of the virus propagation, the shorter it is the faster the peak probability is reached. Sweden, the country, which adopted herd immunization, displays the largest values of t_max_ and Lyapunov time thus the longest time duration for the pandemic. The value of the Lyapunov time decreases depending on the efficacy of the quarantine and other factors (population density etc..). Interestingly enough two almost parallel lines for t_crit_ can be seen the lowest near 50 days and the other below 100 days, the corresponding countries are reported in table AIII. These values are sensitive to the beginning times of the quarantine in each country. A rather random increase of t_max_ may be noticed and attributed to early reopening in different countries. This result supports the first report of the virus spread at the end of 2019 with a possible error of 2 weeks at most [21].

**Figure 13.**
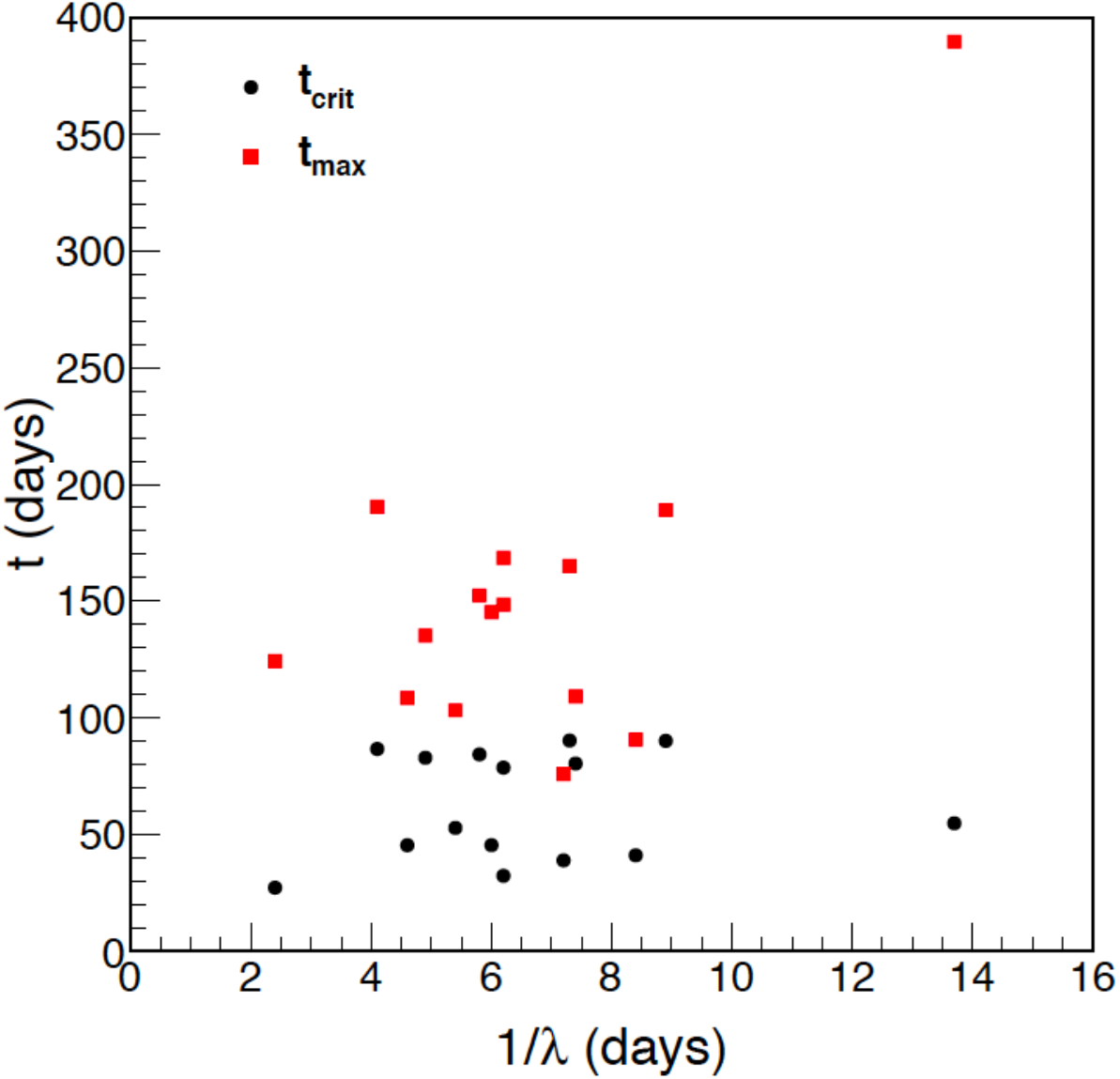
t_max_ & t_crit_ vs 1/λ for all cases analyzed in this paper. The largest values of t_max_ &1/λ refer to Sweden while the largest value of t_crit_ refers to the USA. All the numerical values of this figure are reported in table AIII.

In conclusion, in this paper we have discussed the predictive power of a two-step model based on chaos theory. A comparison among different countries suggests that it would be safe to release the quarantine when the probability for positive is lower than 4%, the maximum value for S. Korea. This implies that, if the quarantine is dismissed, then the same measures, as for the Koreans, should be followed by the other countries: careful testing, backtracking and isolation of positives and quarantine again if needed. Herd immunization or natural selection is very difficult to justify from the data available so far, especially since we are dealing with thousands of human

lives no matter the age or other nonsense. No real dependence on seasonal temperatures is observed, maybe with the only exception of very low temperatures, below 10^°^ C. The model suggests that the beginning time for the pandemic spread is at most as early as the middle of December 2019.

## Data Availability

data available on request from the authors

## APPENDIX Updated data

In this appendix we update some of the relevant figures discussed in the text to the end of June 2020.

**Figure A1.**
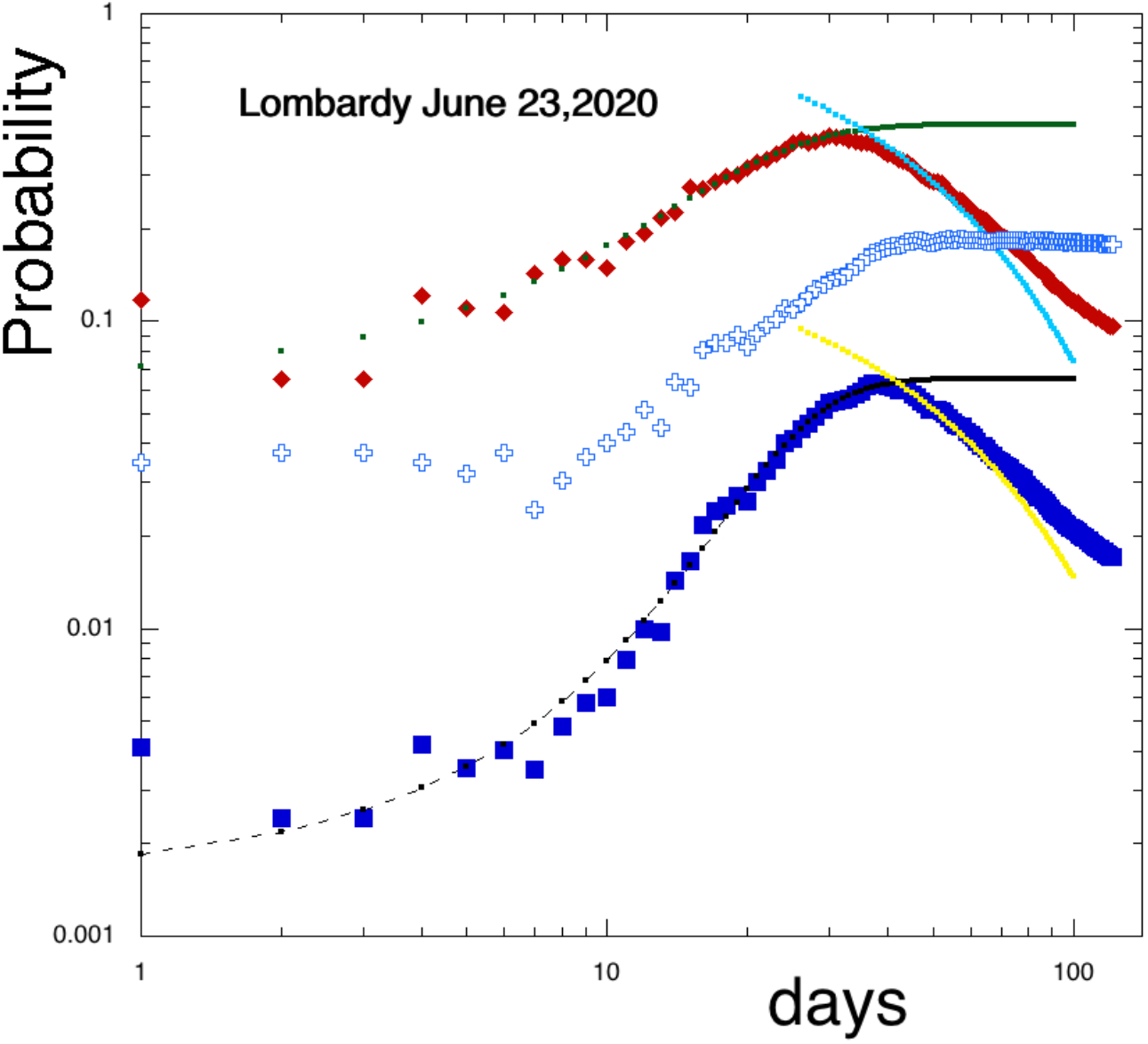
Same as figure 4 for the Lombardy case. The original model predictions are given by the full lines. Notice the data increase respect to the prediction at later times due to the reopening of normal activities: a situation to monitor attentively.

**Figure A2.**
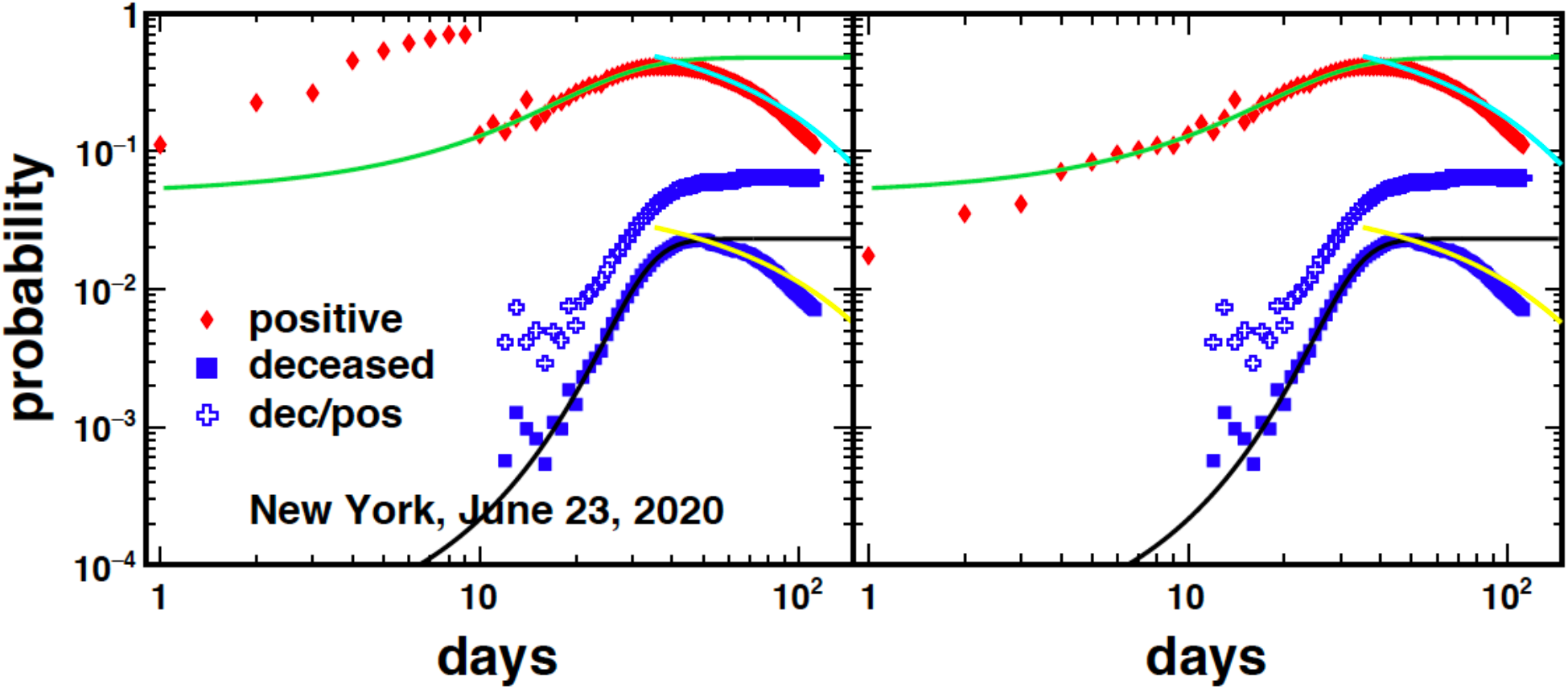
Same as figure 8 updated to June 23,2020. A small decrease respect to the prediction from eq.(1) and (2) is observed at later times.

**Figure A3.**
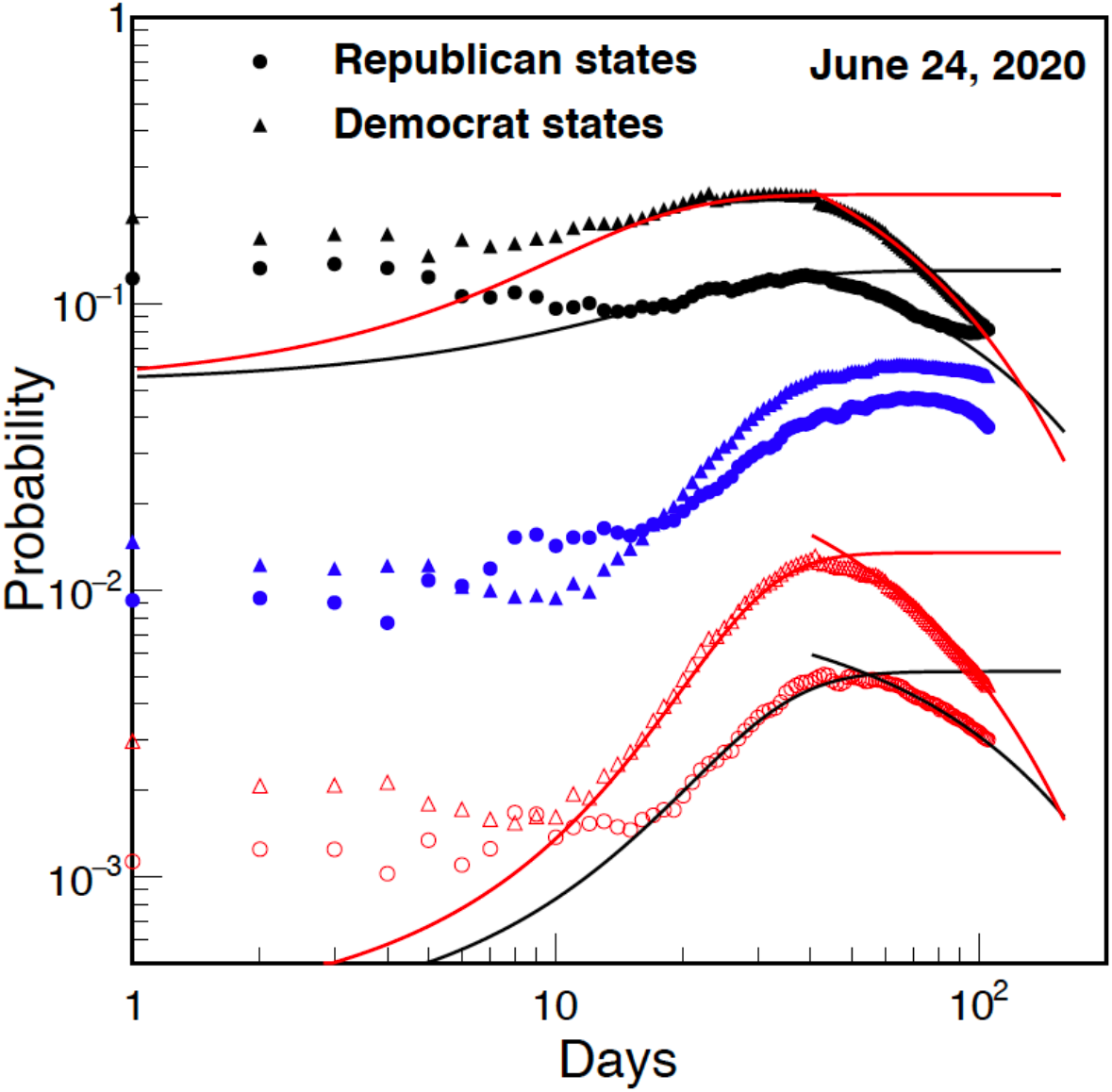
Positive (full symbols) and deceased (open symbols) probabilities for US states with different party governor. The democratic case is dominated by the high population density state of New York, see figure 8. The striking different behavior explains the prediction discussed in fig.10 regarding the USA. Notice an increase at later times for the republican states suggesting a too early reopening. Recall that there is a time delay for the deceased respect to positive.

**Figure A4.**
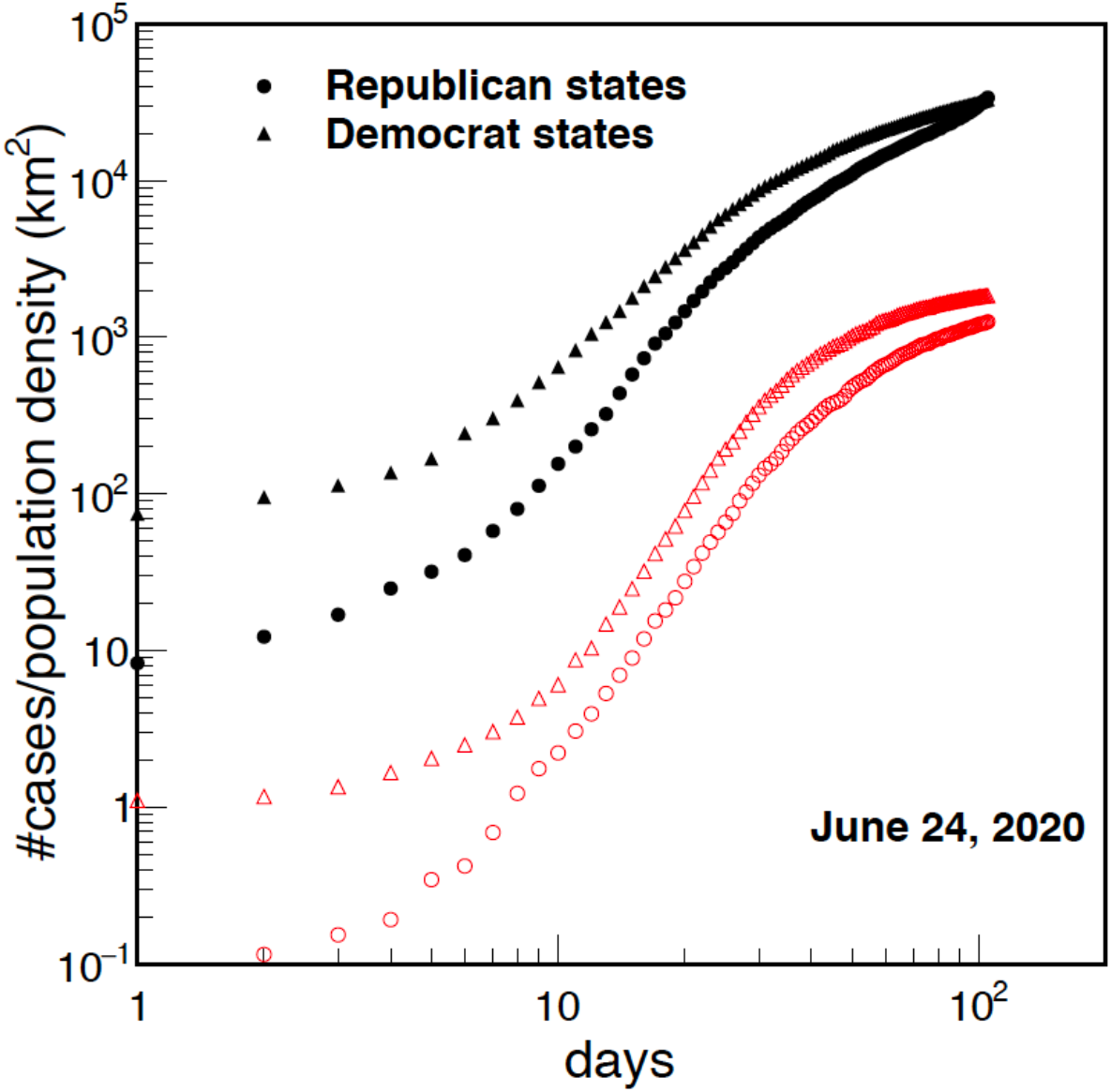
Same as figure A3 but for the number of cases divided the population density, compare to figure 2.

**Figure A5.**
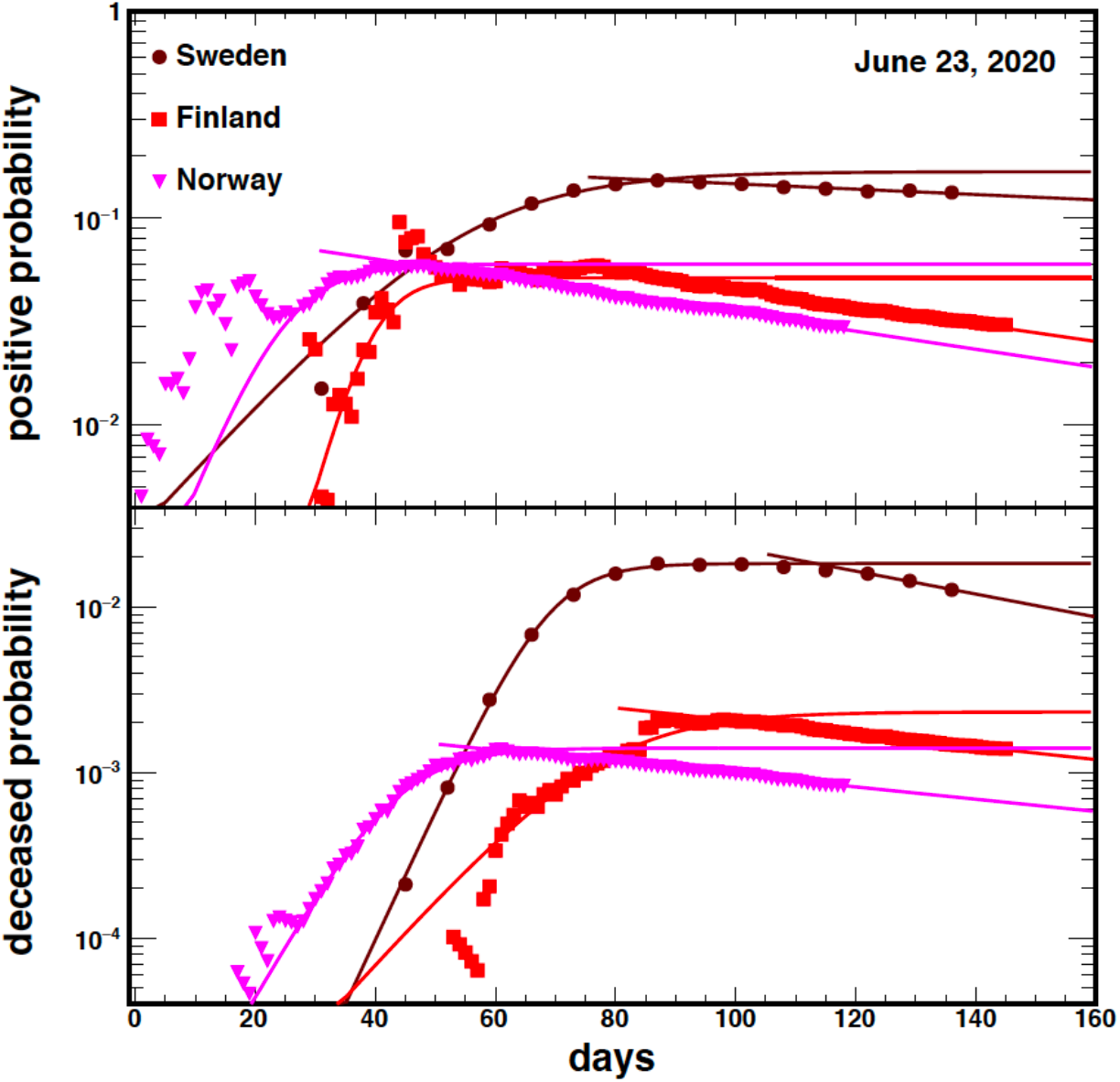
Same as figure 11. Notice the slow decay for Sweden. The data points for Norway have been corrected [*https://ourworldindata.org/grapher/full-list-total-tests-for-covid-19*], compare to fig.11. The updated predictions are given in the table AII.

**Table AI.** Model predictions compared to data for different countries corresponding to figure 10. The France discrepancy is discussed in the text.

| Country | Positive | Positive (quarantine) | Data May 6 | Data June 28 | Deceased | Deceased (quarantine) | Data May 6 | Data June 28 |
| --- | --- | --- | --- | --- | --- | --- | --- | --- |
| S. Korea | $39855 \pm 5978$ | $10880 \pm 1632$ | 10806 | 12715 | $479 \pm 72$ | $292 \pm 44$ | 255 | 282 |
| Japan | $67305 \pm 10096$ | $23457 \pm 3519$ | 15354 | 18390 | $4332 \pm 650$ | $708 \pm 106$ | 543 | 971 |
| Germany | $555076 \pm 83261$ | $189479 \pm 28422$ | 164897 | 193499 | $19350 \pm 2902$ | $10615 \pm 1592$ | 6996 | 8957 |
| Italy | $1994297 \pm 299145$ | $250188 \pm 37528$ | 213013 | 240136 | $188467 \pm 28270$ | $40351 \pm 6053$ | 29315 | 34716 |
| France | $1055857 \pm 158379$ | $457352 \pm 68603$ | 131292 | 156156 | $209604 \pm 31441$ | $114135 \pm 17120$ | 25491 | 29700 |
| New York | $1418146 \pm 212721$ | $398913 \pm 59837$ | 323978 | 392539 | $68931 \pm 10340$ | $26361 \pm 3954$ | 19877 | 24835 |
| Texas | $181520 \pm 27228$ | $55075 \pm 8261$ | 34422 | 148728 | $5518 \pm 828$ | $2623 \pm 393$ | 948 | 2393 |
| USA | $4490440 \pm 673566$ | $1487103 \pm 223065$ | 1171185 | 2452048 | $260530 \pm 39080$ | $87963 \pm 13194$ | 68081 | 124811 |
| UK | $3164339 \pm 474651$ | $287538 \pm 43131$ | 194994 | 310254 | $433073 \pm 64961$ | $45713 \pm 6857$ | 29427 | 43514 |

**Table AII.** Updated results on June 23,2020 for Sweden, Norway and Finland. Notice that the discrepancy between Norway and Finland discussed in the main text was due to a mistake in the data reporting of Norway [*https://ourworldindata.org/grapher/full-list-total-tests-for-covid-19]*

| Country | Positive-model | Data Positive | Deceased-model | Data Deceased |
| --- | --- | --- | --- | --- |
| Sweden | 58270±8740 | 65137 | 4730±710 | 5280 |
| Finland | 7331±1100 | 7191 | 338±51 | 328 |
| Norway | 8518±1278 | 8815 | 242±36 | 249 |

**Table AIII.** Typical times obtained from the model fits to data for different countries corresponding to figure 13.

| Country (region) | $1/\lambda$ (days) | $t_{crit}$(days) | $t_{max}$(days) | Population density ($km^{-2}$) |
| --- | --- | --- | --- | --- |
| S. Korea | 2.4 | 27.3 | 124.2 | 511.6 |
| Japan | 8.9 | 90.2 | 189.0 | 334.7 |
| Germany | 6.2 | 78.7 | 168.6 | 234.6 |
| Italy | 5.4 | 52.9 | 103.3 | 200.6 |
| France | 5.8 | 84.4 | 152.4 | 118.3 |
| New York | 8.4 | 41.2 | 90.8 | 137.6 |
| Texas | 6.2 | 32.4 | 148.5 | 42.4 |
| USA | 7.3 | 90.3 | 165.1 | 36.2 |
| UK | 7.4 | 80.5 | 109.3 | 279.5 |
| Sweden | 13.7 | 54.9 | 389.6 | 24.6 |
| Finland | 4.1 | 86.7 | 190.4 | 18.2 |
| Norway | 6.0 | 45.5 | 145.3 | 14.8 |
| Florida | 4.6 | 45.5 | 108.6 | 129.1 |
| Lombardy | 7.2 | 39.0 | 76.0 | 421.7 |
| Spain | 4.9 | 82.9 | 135.3 | 92.4 |

